# Sociodemographic Determinants of Respiratory Failure in Patients Hospitalized with SARI in Cali, Colombia

**DOI:** 10.1101/2025.04.08.25325483

**Authors:** Pablo Roa-Urrutia, Andrés Felipe Valencia-Cardona, Carlos Reina, Gissel Guzmán-Escarria, Jorge Iván Montoya-Salazar, Anthony Garcés-Hurtado, María Eugenia Ortiz-Carrillo, David Arango-Londoño, German Ávila, Sebastian Cruz Barbosa, Alberto Concha-Eastman

## Abstract

**Objectives:** To analyze the association between respiratory failure and sociodemographic determinants in patients hospitalized with Severe Acute Respiratory Infection (SARI) in Cali, Colombia, in 2022.

**Method:** A descriptive and cross-sectional study was conducted using data from the 2022 respiratory failure notification file No. 348. A generalized linear model with an exponential binomial family was applied. Absolute frequencies, proportions, and odds ratios (OR) were estimated.

**Results:** Of 1,163 patients, 4.5% (n=52) developed respiratory failure. Those with high [OR: 2.03; 95%CI: 1.2–2.8] or medium-high [OR: 1.4; 95%CI: 0.9–3.7] vulnerability had a greater likelihood of respiratory failure compared to patients with low vulnerability. Asthma and COPD increased the risk [OR: 4.3; 95%CI: 1.5–6.03] and [OR: 3.3; 95%CI: 1.3–4.3], respectively. Sepsis was also identified as a significant risk factor [OR: 2.7; 95%CI: 1.8–3.5].

**Conclusions:** The results emphasize the need for targeted public health actions. Sociodemographic factors, especially multidimensional poverty such as limited education, income, and healthcare access play a key role in respiratory failure outcomes.

## INTRODUCTION

Severe Acute Respiratory Infection (SARI) are a significant contributor to morbidity, encompassing a spectrum of severity that can range from moderate to severe, potentially resulting in deadly consequences. The World Health Organization (WHO) identifies several infectious agents with pandemic potential, including influenza, syncytial virus, adenovirus, and SARS-CoV-2 (1).

SARIs encompass a range of clinical characteristics that have implications for community health, specifically targeting the respiratory tract, particularly the lower respiratory tract. The aforementioned symptoms arise due to the presence of infectious organisms, encompassing a wide range of diseases caused by bacteria, viruses, parasites, or fungi. Some of the most prevalent symptoms include coughing, respiratory failure, and dyspnea, among other manifestations (1).

At the 15th meeting of the International Health Regulations Emergency Committee in May 2023, WHO said that COVID-19 is now a known problem and is no longer a public health emergency of international concern (PHEIC). As a result, its strategic preparedness and response plan for 2023–2025 has been updated (2).

On June 30, 2022, the COVID-19 health emergency in Colombia was officially terminated by the Ministry of Health and Social Protection, in accordance with Resolution 1705 of 2023 (3), making the disease part of the cluster of infectious agents monitored in the country by the National Institute of Health (Instituto Nacional de Salud-INS) (1).

Pan American Health Organization (PAHO) reported that as of efpidemiological week (EW) 43 of 2022, SARIs, measured by hospitalizations, were low compared to the historical figure of the last 5 years in Latin America and the Caribbean, except for Bolivia where the intensity was moderate, with an increase in influenza since EW 36 in Argentina, Brazil, Uruguay, Chile, and Paraguay. Influenza A(H3N2) predominated, with A(H1N1) pdm09 and B/Victoria also circulating. Colombia, Peru, Ecuador, Bolivia, and Venezuela showed modest respiratory syncytial virus activity (1).

In Latin America, the burden of SARIs has varied over time. In Colombia, SARIs accounted for 7,796,644 outpatient and emergency visits in 2022, with 294,408 hospitalizations (4).

As a reference center for the entire southwestern region of Colombia, the city of Cali, a major urban center with 2.4 million residents, serves as a regional referral hub for respiratory illnesses (5). The National Public Health Surveillance System (SIVIGILA) monitors SARIs, assessing their epidemiological and sociodemographic impact. Because it enables the characterization of prevalent and stationary etiological agents as well as the assessment of their behavior and sociodemographic and health features in this region of Colombia, public health and epidemiological surveillance of SARIs in this city are therefore important. This in turn encourages alarm systems for the creation of plans and the distribution of mitigation, control, and prevention actions (6,7).

As part of the national surveillance of SARIs in 2022, respiratory syncytial virus made up 34% of the agents found. It was followed by adenovirus with 21.8%, rhinovirus with 7.7%, and influenza type A with a predominant subtype A (H3N2) with 6.4%. For every case reported to the Cali monitoring system by municipality of residence in 2022, respiratory syncytial virus made up 47%, adenovirus made up 19%, and influenza A (H3N2) made up 12.5% (1).

As indicated, public health monitoring also assesses processes connected with the sociodemographic characteristics of the population to recognize groups at risk of severe morbidity. This can be determined by looking at factors outside of medical care that influence patients’ health, such as their environment and lifestyle (1). The multidimensional poverty index (MPI) should also be included because it affects people’s health in several ways. It takes a holistic view by measuring the space between its constituent indicators. First, a profile of each person and family’s level of deprivation is created. The indicators used to measure deprivation cover areas including health, education, and income (8).

Based on the aforementioned considerations, the lack of comprehensive research pertaining to epidemiological surveillance and public health regarding sociodemographic and health factors that may contribute to severe morbidities resulting from Severe Acute Respiratory Infection (SARI), specifically respiratory failure, creates an opportunity for studies such as the present one. The objective was to estimate the prevalence rates and examine the associations between the risk of hospital admissions for SARI and the occurrence of respiratory failure due to sociodemographic and health-related factors in the city of Cali, Colombia, throughout the year 2022.

## MATERIAL AND METHOD

This study employed a descriptive and cross-sectional analytical approach, allowing for the characterization and analysis of associations within the sample. This design enables the identification of event frequency, population-level patterns, and relationships between exposure and outcome. This study was based on a retrospective review of the records maintained by the Secretariat of Health of Cali, specifically notification file 348 of 2022, which is registered in the National Public Health Surveillance System (Sistema de Vigilancia en Salud Pública-SIVIGILA). This file reports unusual Severe Acute Respiratory Infection (SARI) cases based on contact history, travel, atypical severity, or treatment failure. Cases are classified using clinical, epidemiological, and laboratory data. Considering that the INS closes annual information in March of the following year, the database for 2022 was considered; thus, the most recent data accessible for this inquiry pertains to the 2022 cohort, as the closing date for annual information was March 2023. Additionally, on June 30, 2022, the Colombian Ministry of Health declared the end of the COVID-19 pandemic, marking a suitable date to exclude SARI cases linked to COVID-19 (3).

Since the data are derived from the city’s population registry, the implementation of a sampling size calculation and design was unnecessary. The inclusion criteria encompassed all individuals who were recorded in notification file 348 with respect to hospital admissions in 2022. Patients with inconsistent data in notification file 348, those who had been notified multiple times in the SIVIGILA information system, and those who had contracted COVID-19 as an infectious agent were excluded. Following the completion of the data quality and criteria processing, a final sample of 1.163 observations was obtained from the 1.204 notifications, of which 41 were inconsistent values (3.4 %).

The dependent variable was respiratory failure as defined by the National Heart, Lung, and Blood Institute. Respiratory failure is a critical condition that substantially compromises an individual’s capacity to breathe independently (9), attaining a binary result (exhibiting respiratory failure or lacking it). The categorical covariates included age group, sex, and affiliation regime, as well as the multidimensional poverty index (MPI), which was calculated in the geographic unit per block and was established by the National Administrative Department of Statistics (Departamento

Administrativo Nacional de Estadística-DANE) at the national level. However, for the purposes of this study, an intersection was performed between the block shapefile and the neighborhood to determine the average MPI per neighborhood. Furthermore, by subdividing the aforementioned averages into quintiles, the vulnerability categories (Low Vulnerability, Medium-Low Vulnerability, Medium Vulnerability, Medium-High Vulnerability, and High Vulnerability) were ascertained. Patients with immunosuppressed conditions (cancer and renal failure), comorbidities (including asthma, chronic obstructive pulmonary disease (COPD), diabetes, heart disease, and hypertension), and respiratory failure-related complications (including sepsis, pleural effusion, and pericardial effusion).

The proposed variables and their relationship are considered in accordance with the literature’s recommendations and the available data sources. Similarly, correlation between the subjects’ observations is avoided in this type of research by considering only the initial report of respiratory failure.

The present study lacks individual-level data and does not involve any form of intervention with the participants. The study’s findings were derived through the estimation of absolute and relative frequencies and proportions. The significance level for the comparative analysis, conducted using the chi-square (χ^2^) independence test, was established at 0.05 (10). The statistical study of several variables using a Generalized Linear Model (GLM) with a canonical logic link. Next, a progressive selection was performed using p-values less than 0.20 (Stepwaise) and elements that increase the Akaike Information Criterion (AIC) for regressor variable selection were integrated. The regression coefficient exponents were used to construct the Odds Ratio (OR) to assess the association (11). The Global Likelihood-Ratio analysis revealed a significant p-value of < 0.01, indicating a linear relationship between variables and outcomes. Patients with respiratory failure had a sensitivity of 78.7%. Finally, 11 data were found with a leverage effect from residuals that may be affecting the coefficients, but we kept the model because the significance and estimates weren’t affected. Geographic characteristics of the city’s urban area at the neighborhood level were used to describe each region. It was limited to January 1–December 31, 2022, using SIVIGILA notification file 348 and the Cali Mayor’s Office’s official planning cartography basis (12).

All of the above methods were carried out through the specialized statistical program in data science in Rstudio (version 4.4.3, R Foundation for Statistical Computing, Boston, USA) (13) y QGis Desktop 3.18.3 where the shapefile layer for spatial analysis was obtained (14). The study obtained ethical approval from the Ethics Committee of the district public health secretariat of Cali, Colombia, Record No. 202441450100041674.

## RESULTS

### Descriptive analysis by outcome

In 2022, 4.5% of cases reported by epidemiological surveillance showed unusual ARIs in the city of Cali. These people were hospitalized and had respiratory failure. The age group with the highest rate of respiratory failure is children under 18 years (45.5%), followed by women (52.8%) and men (67.9%). In MPI, the groups with the highest rates of respiratory failure were those with medium-high vulnerability (30.2%) and high vulnerability (22.6%) (see Table 1). People who had asthma, COPD, diabetes, heart disease, or hypertension were 11.3%, 17%, 9.4%, 9.4%, and 9.4% more likely to have respiratory failure. Cancer patients whose immune systems have declined had 3.8% and 5.7% of them had renal failure (see Table 1). 7.5% of people with sepsis had complications related to breathing failure, 7.5% had pleural effusion, and 1.9% had pericardial effusion (see Table 1). The variables considered significant for this study were MPI (p< 0.05), asthma (p< 0.05), COPD (p< 0.05) and sepsis (p< 0.001) (see Table 1).

**Table 1.**
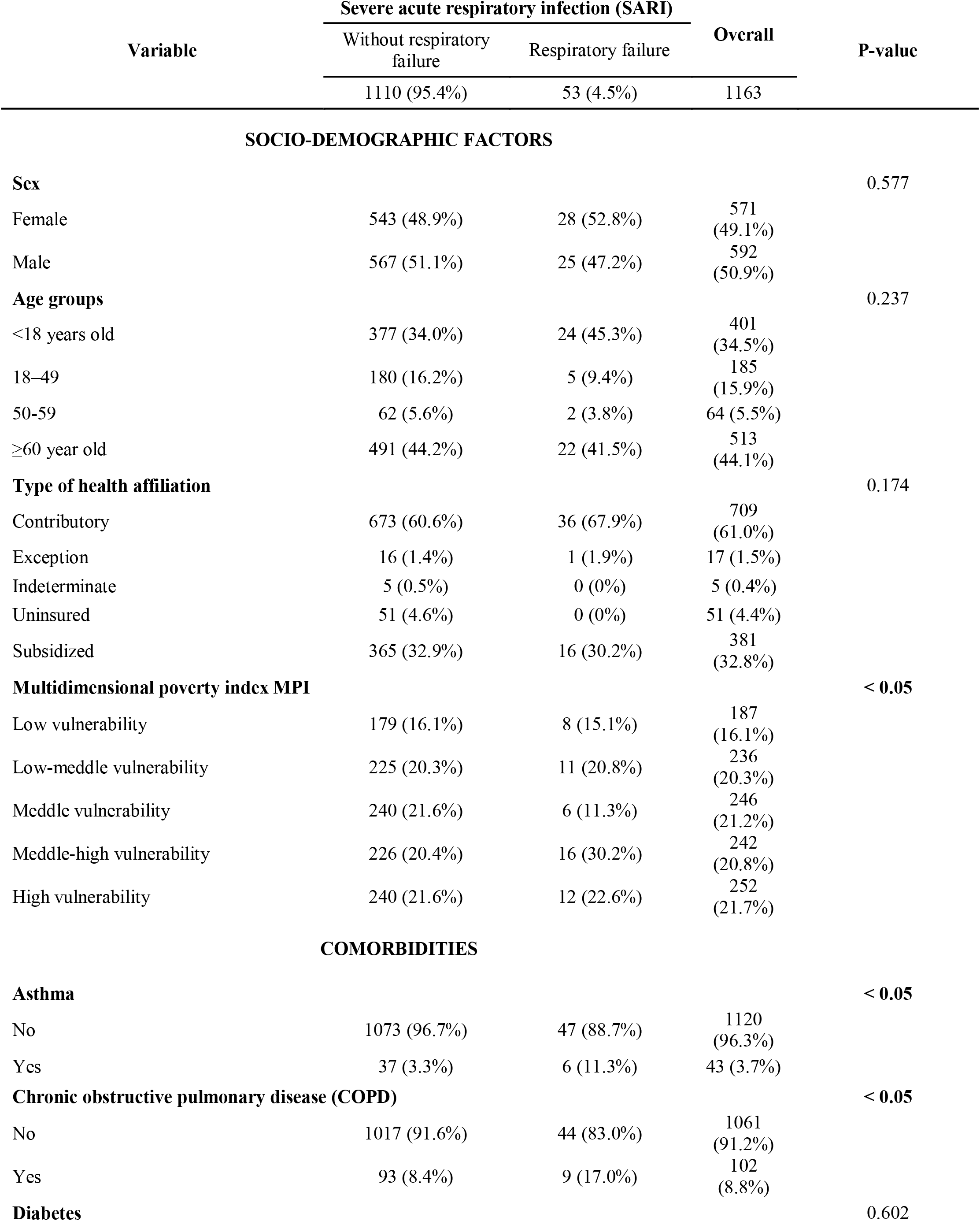
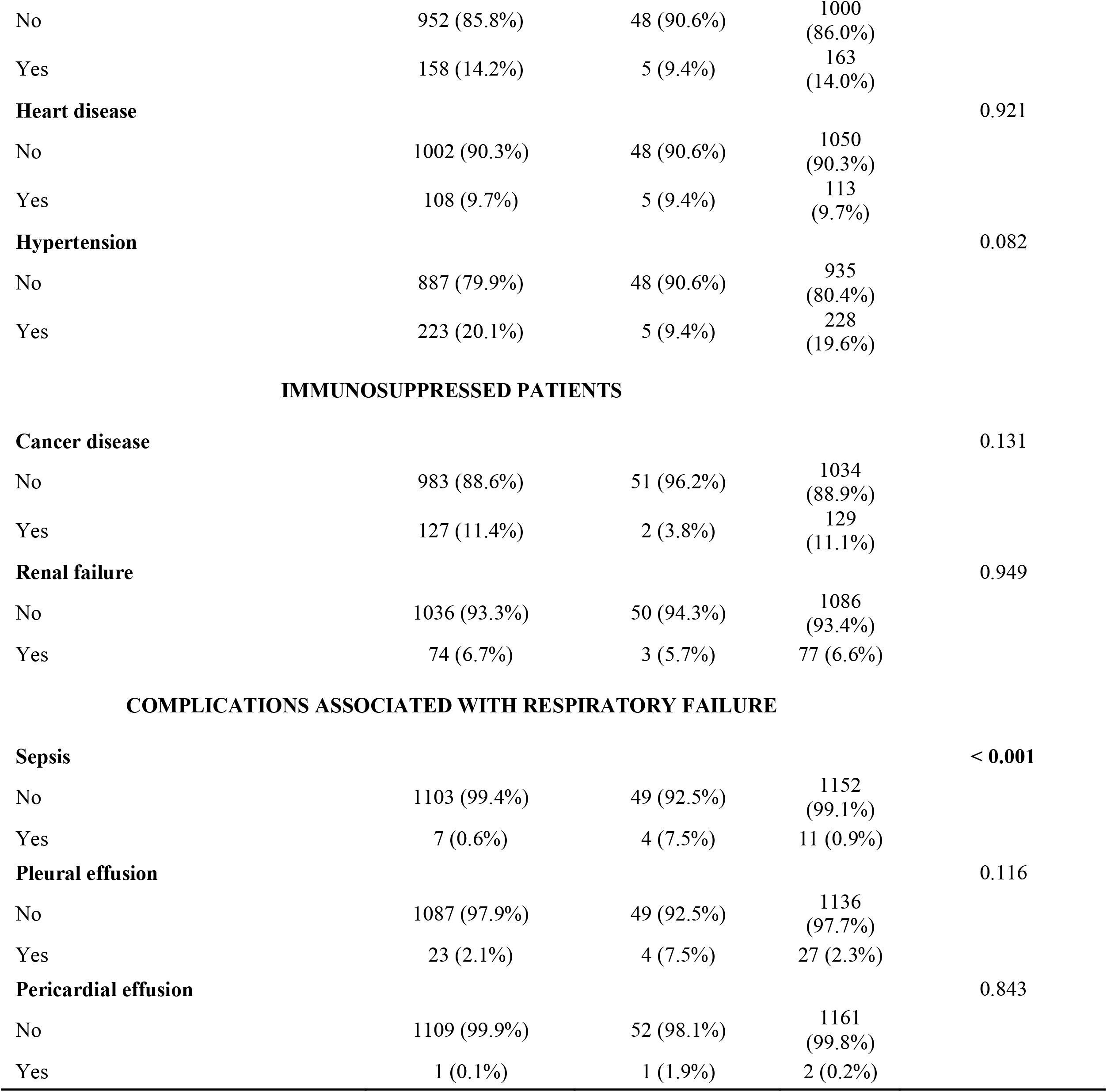
Descriptive analysis and χ^2^ chi-square independence test of respiratory failure in 2022, Cali, Colombia

### Inferential analysis by outcome

The final GLM for the IMP variable showed that people with a high [OR: 2.03 (95%CI: 1.2–2.8)] or medium-high [OR: 1.4 (95%CI: 0.9–3.7)] vulnerability might be more likely to have respiratory failure than people with a low vulnerability. Another important thing to note is that the asthma variables had a higher odds ratio than those who did not have this comorbidity when it came to the section on comorbidities and complications associated with respiratory failure [OR: 4.3 (95%CI: 1.5–6.03)]. Similarly, for patients with COPD [OR: 3.3 (95%CI: 1.3–4.3)], there was an increased risk of respiratory failure compared with people without COPD. Finally, sepsis was estimated to be a risk factor for respiratory failure [OR: 2.7 (95%CI: 1.8–3.5)], since the risk was almost 3 times higher compared to those who did not have this complication (see Table 2).

**Table 2.**
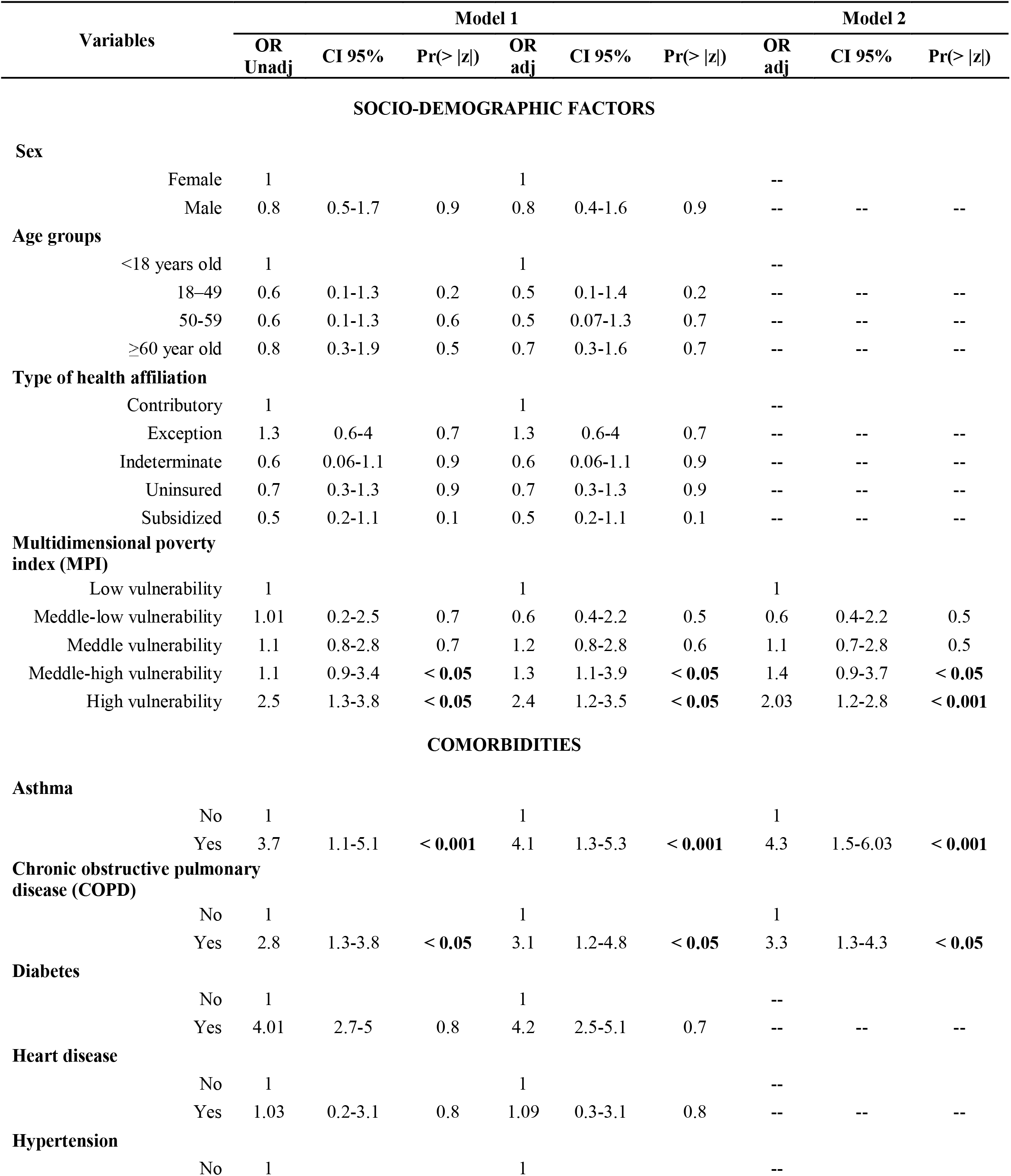
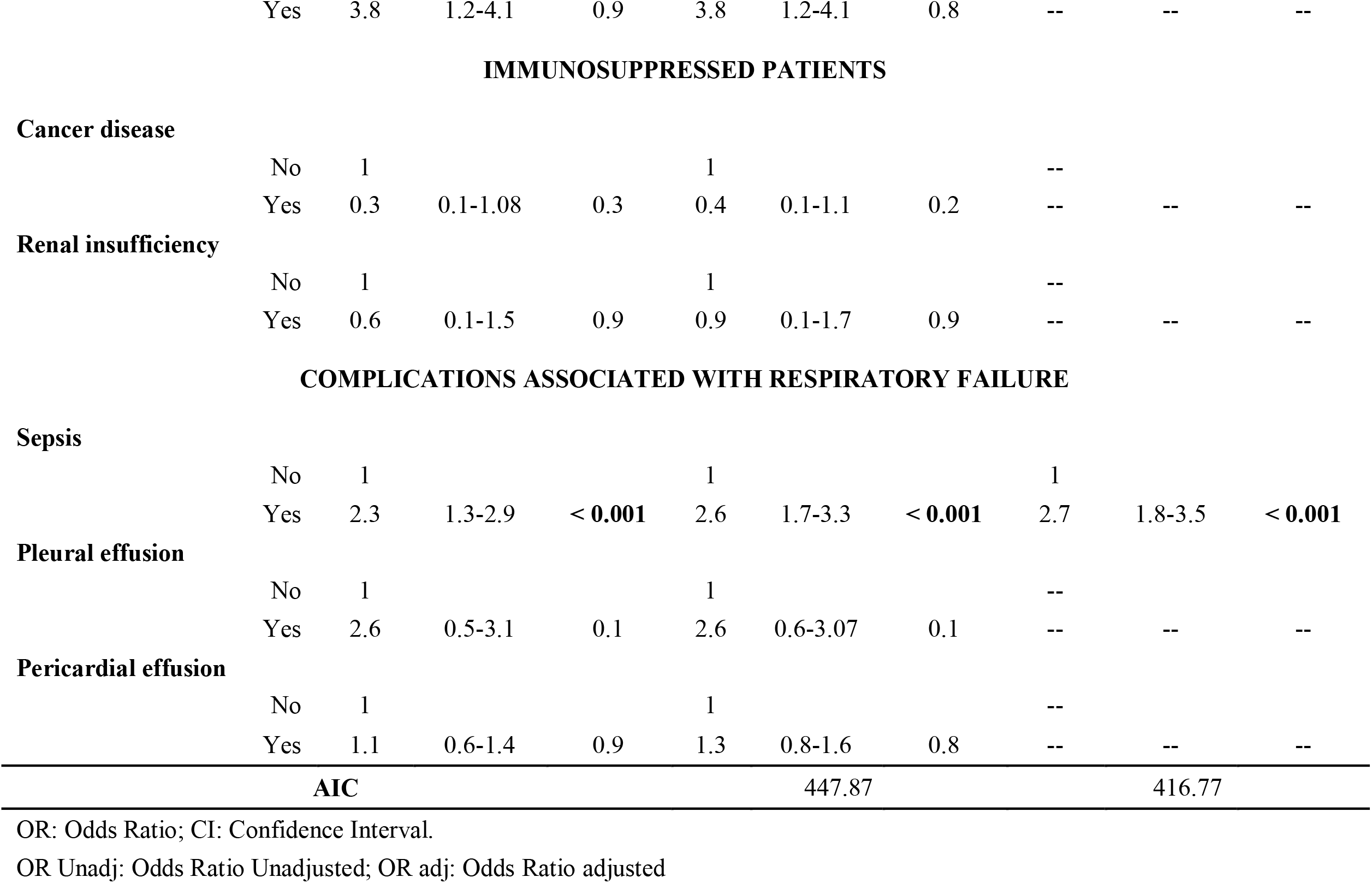
Inferential Analysis, General Linear Model, and Wald Test of respiratory failure in 2022, Cali, Colombia

| Variables | Model 1 |  |  |  |  |  | Model 2 |  |  |
| --- | --- | --- | --- | --- | --- | --- | --- | --- | --- |
|  | OR Unadj | CI 95% | Pr(> \|z\|) | OR adj | CI 95% | Pr(> \|z\|) | OR adj | CI 95% | Pr(> \|z\|) |
| **SOCIO-DEMOGRAPHIC FACTORS** |  |  |  |  |  |  |  |  |  |
| **Sex** |  |  |  |  |  |  |  |  |  |
| Female | 1 |  |  | 1 |  |  | -- |  |  |
| Male | 0.8 | 0.5-1.7 | 0.9 | 0.8 | 0.4-1.6 | 0.9 | -- | -- | -- |
| **Age groups** |  |  |  |  |  |  |  |  |  |
| <18 years old | 1 |  |  | 1 |  |  | -- |  |  |
| 18–49 | 0.6 | 0.1-1.3 | 0.2 | 0.5 | 0.1-1.4 | 0.2 | -- | -- | -- |
| 50-59 | 0.6 | 0.1-1.3 | 0.6 | 0.5 | 0.07-1.3 | 0.7 | -- | -- | -- |
| ≥60 year old | 0.8 | 0.3-1.9 | 0.5 | 0.7 | 0.3-1.6 | 0.7 | -- | -- | -- |
| **Type of health affiliation** |  |  |  |  |  |  |  |  |  |
| Contributory | 1 |  |  | 1 |  |  | -- |  |  |
| Exception | 1.3 | 0.6-4 | 0.7 | 1.3 | 0.6-4 | 0.7 | -- | -- | -- |
| Indeterminate | 0.6 | 0.06-1.1 | 0.9 | 0.6 | 0.06-1.1 | 0.9 | -- | -- | -- |
| Uninsured | 0.7 | 0.3-1.3 | 0.9 | 0.7 | 0.3-1.3 | 0.9 | -- | -- | -- |
| Subsidized | 0.5 | 0.2-1.1 | 0.1 | 0.5 | 0.2-1.1 | 0.1 | -- | -- | -- |
| **Multidimensional poverty index (MPI)** |  |  |  |  |  |  |  |  |  |
| Low vulnerability | 1 |  |  | 1 |  |  | 1 |  |  |
| Meddle-low vulnerability | 1.01 | 0.2-2.5 | 0.7 | 0.6 | 0.4-2.2 | 0.5 | 0.6 | 0.4-2.2 | 0.5 |
| Meddle vulnerability | 1.1 | 0.8-2.8 | 0.7 | 1.2 | 0.8-2.8 | 0.6 | 1.1 | 0.7-2.8 | 0.5 |
| Meddle-high vulnerability | 1.1 | 0.9-3.4 | < **0.05** | 1.3 | 1.1-3.9 | < **0.05** | 1.4 | 0.9-3.7 | < **0.05** |
| High vulnerability | 2.5 | 1.3-3.8 | < **0.05** | 2.4 | 1.2-3.5 | < **0.05** | 2.03 | 1.2-2.8 | < **0.001** |
| **COMORBIDITIES** |  |  |  |  |  |  |  |  |  |
| **Asthma** |  |  |  |  |  |  |  |  |  |
| No | 1 |  |  | 1 |  |  | 1 |  |  |
| Yes | 3.7 | 1.1-5.1 | < **0.001** | 4.1 | 1.3-5.3 | < **0.001** | 4.3 | 1.5-6.03 | < **0.001** |
| **Chronic obstructive pulmonary disease (COPD)** |  |  |  |  |  |  |  |  |  |
| No | 1 |  |  | 1 |  |  | 1 |  |  |
| Yes | 2.8 | 1.3-3.8 | < **0.05** | 3.1 | 1.2-4.8 | < **0.05** | 3.3 | 1.3-4.3 | < **0.05** |
| **Diabetes** |  |  |  |  |  |  |  |  |  |
| No | 1 |  |  | 1 |  |  | -- |  |  |
| Yes | 4.01 | 2.7-5 | 0.8 | 4.2 | 2.5-5.1 | 0.7 | -- | -- | -- |
| **Heart disease** |  |  |  |  |  |  |  |  |  |
| No | 1 |  |  | 1 |  |  | -- |  |  |
| Yes | 1.03 | 0.2-3.1 | 0.8 | 1.09 | 0.3-3.1 | 0.8 | -- | -- | -- |
| **Hypertension** |  |  |  |  |  |  |  |  |  |
| No | 1 |  |  | 1 |  |  | -- |  |  |
| Yes | 3.8 | 1.2-4.1 | 0.9 | 3.8 | 1.2-4.1 | 0.8 | -- | -- | -- |
| Cancer disease |  |  |  |  |  |  |  |  |  |
| No | 1 |  |  | 1 |  |  | -- |  |  |
| Yes | 0.3 | 0.1-1.08 | 0.3 | 0.4 | 0.1-1.1 | 0.2 | -- | -- | -- |
| Renal insufficiency |  |  |  |  |  |  |  |  |  |
| No | 1 |  |  | 1 |  |  | -- |  |  |
| Yes | 0.6 | 0.1-1.5 | 0.9 | 0.9 | 0.1-1.7 | 0.9 | -- | -- | -- |
| Sepsis |  |  |  |  |  |  |  |  |  |
| No | 1 |  |  | 1 |  |  | 1 |  |  |
| Yes | 2.3 | 1.3-2.9 | < 0.001 | 2.6 | 1.7-3.3 | < 0.001 | 2.7 | 1.8-3.5 | < 0.001 |
| Pleural effusion |  |  |  |  |  |  |  |  |  |
| No | 1 |  |  | 1 |  |  | -- |  |  |
| Yes | 2.6 | 0.5-3.1 | 0.1 | 2.6 | 0.6-3.07 | 0.1 | -- | -- | -- |
| Pericardial effusion |  |  |  |  |  |  |  |  |  |
| No | 1 |  |  | 1 |  |  | -- |  |  |
| Yes | 1.1 | 0.6-1.4 | 0.9 | 1.3 | 0.8-1.6 | 0.8 | -- | -- | -- |

|  |  |  |
| --- | --- | --- |
| AIC | 447.87 | 416.77 |

OR: Odds Ratio; CI: Confidence Interval.
OR Unadj: Odds Ratio Unadjusted; OR adj: Odds Ratio adjusted

### Descriptive analysis by territory

Aspects of interest such as the northern zone are highlighted in the neighborhood rates of SARIs; among these, the Ciudadela Floralia code 0610 (meddle-high vulnerability) neighborhood has the greatest rate at 1.08 cases per 100 000 inhabitants (see map 1).

With a vulnerability rating of 0.68, the Ciudad Córdoba neighborhood code 1596 is in commune 15, in the eastern zone. Ciudad 2000 code 1697 (meddle-low vulnerability) and Antonio Nario code 1604 (high vulnerability), both with a rate of 0.68, are among the localities in commune 16 that exhibit high vulnerability.

In commune 17, particularly in the southern zone, low vulnerability localities stand out, including El Ingenio code 1780 (0.59) and Lili code 1775 (0.72), as well as Mayapan-Las Vegas code 1781 (0.68), which has a meddle-low vulnerability rating. Furthermore, within the central zone, the rates of cases per 100 000 inhabitants are 0.59 in San Nicolás code 0312 (meddle-high vulnerability) in Comuna 3 and 0.59 in El Guabal code 1002 (meddle-high vulnerability) in Comuna 10 (see map I).

## DISCUSSION

The results of this study indicate that patients with SARI who have a high or meddle-high MPI are at a greater risk for respiratory failure than those with a low MPI. Similarly, the descriptive analysis by territory revealed that neighborhoods with a high or meddle-high MPI had the greatest rate of respiratory failure. According to the literature, indicates that populations characterized by a high MPI are more susceptible to experiencing health issues. This is exemplified in the city of Cúcuta, Colombia, where a proportional rate of SARI among migrants and individuals under the age of 15 increased by one unit. A similar relationship was observed between high IMP and increases in SARI rates of 1.25 and 1.08, respectively (15). In another country like India, they found that people with high multidimensional poverty were more likely to have Tuberculosis (TB) [OR: 1.82 (95%CI: 1.73–1.90)] compared to non-poor people (16). In India, they found that people with high multidimensional poverty were more likely to have TB [OR: 1.82 (95%CI: 1.73–1.90)] compared to non-poor people (17).

Based on information gathered by the Centers for Disease Control and Prevention (CDC), it has been determined that around 10% of the global population is afflicted with asthma. Moreover, there has been an approximate 15% rise in the prevalence of this condition in the United States over the course of the last two decades. An overall of 5% of the individuals under consideration are categorized as having severe asthma.

The prevalence of respiratory failure necessitating ventilatory assistance among hospitalized adult patients with asthma is reported to range from 3% to 16%. A mortality rate of around 10% has been documented among patients who are admitted to the intensive care unit (ICU) with asthmatic condition (18).

It is worth mentioning that COPD patients diagnosed with SARIs had a greater risk of developing respiratory failure in comparison to those without this comorbidity. This finding is consistent with previous research, which supports the notion that respiratory failure is directly caused by acute airway constriction and a critical increase in airway resistance. Severe exacerbations of COPD are the primary etiologies of respiratory failure. Other risk factors include prior respiratory infections, physical overexertion, environmental pollutant exposure, pre-existing cardiac conditions, non-compliance with treatment, and environmental pollution. As a result, it is possible to induce hypoxemia and hypercapnia, which raises the likelihood of ICU admission requiring invasive or non-invasive ventilation support (19). In Spain, studies found that about one-third of patients who were sent home because of a COPD flare-up are readmitted within three months. These studies found that heart failure and anemia were linked to readmission. The death rate for this group of patients with respiratory failure was also about 7% (20).

Finally, our investigation showed that individuals who came with sepsis had a higher likelihood of experiencing respiratory failure in comparison to those who did not exhibit this associated consequence. Respiratory failure is frequently observed in cases of sepsis, constituting around 40% of the underlying causes. It is noteworthy that a subset of patients with sepsis, ranging from 6 to 7%, may experience respiratory failure. Importantly, this complication can arise from several anatomical sites or distinct viral sources. The use of timely detection and intervention measures for sepsis represents the most effective approach in mitigating the potential for respiratory difficulties that may arise in conjunction with this condition (20).

## LIMITATIONS AND STRENGTHEN

One potential constraint that might have emerged is the failure to report acute respiratory infections affecting patients, as some of those with severe symptoms who originated in Cali could have received treatment in other territorial entities. However, this study may provide information for future seroprevalence investigations even though sentinel surveillance of infectious agents that could contribute to severe respiratory symptoms and respiratory failure was a further limitation. The analyses conducted by territory identified neighborhoods with high and medium-high MPI, which exhibited high percentage rates consistent with the inferential analysis performed via the regression model. Nevertheless, certain neighborhoods with medium-low and low MPI exhibited high rates, indicating the presence of confounding factors in these areas that contributed to these results, which would require additional analysis.

The social determinants of health approach, which proposes a multidimensional view of SARIs that transcends clinical perspectives and helps to identify at-risk individuals based on sociodemographic characteristics, which are then utilized as input for public health decision-making and epidemiological surveillance strategies, is one of the study’s strengths.

## RECOMMENDATIONS

This analysis underscores the significance of addressing these medical conditions within the healthcare sector, considering their clinical manifestations. Additionally, it recognizes the influence of sociodemographic factors, particularly those related to multidimensional poverty, such as educational attainment, financial resources, and limited access to healthcare services, in contributing to adverse health outcomes. Therefore, it is recommended for policymakers to make public health decisions based on evidence of clinical symptoms, comorbidities and sociodemographic factors that adjust to the realities of the territories.

## CONCLUSIONS

This study has revealed facts pertaining to respiratory failure in patients with various health conditions, encompassing those with high and medium-high MPI scores. In conclusion, it is evident that international poverty exerts a significant influence on health outcomes, particularly in relation to respiratory failure. Another noteworthy aspect to consider is the presence of comorbidities such as Asthma and COPD, which might serve as risk factors for respiratory failure due to their impact on the complications and severity of respiratory symptoms. The heightened susceptibility of patients with sepsis to the onset of respiratory failure can be attributed to the prevalence of sepsis as a primary catalyst for respiratory failure. Consequently, the prompt detection and intervention of sepsis are imperative in mitigating the likelihood of respiratory failure.

## Data Availability

All data produced in the present study are available upon reasonable request to the authors

## CONFLICT OF INTEREST

The authors have no conflicts of interest associated with the material presented in this paper.

## ACKNOWLEDGMENT

The authors extend their gratitude to the Secretariat of Health of Cali and all hospital and private insurer health personnel for facilitating prompt treatment of patients with acute respiratory infections in 2022.

## STATEMENT ABOUT AUTHORS’ CONTRIBUTIONS

PR contributed to the topic, design, data analysis, interpretation, discussion, limitations, and strengthening. AV also contributed to the topic through design, interpretation, discussion, limitations, and strengthening. CR, and DA contributed to the topic, design, data analysis and interpretation. GG and GA also contributed to the design, spatial analysis, and interpretation. JM, AG, MO, SC, and AC contributed to the topic, design, and discussion.

---

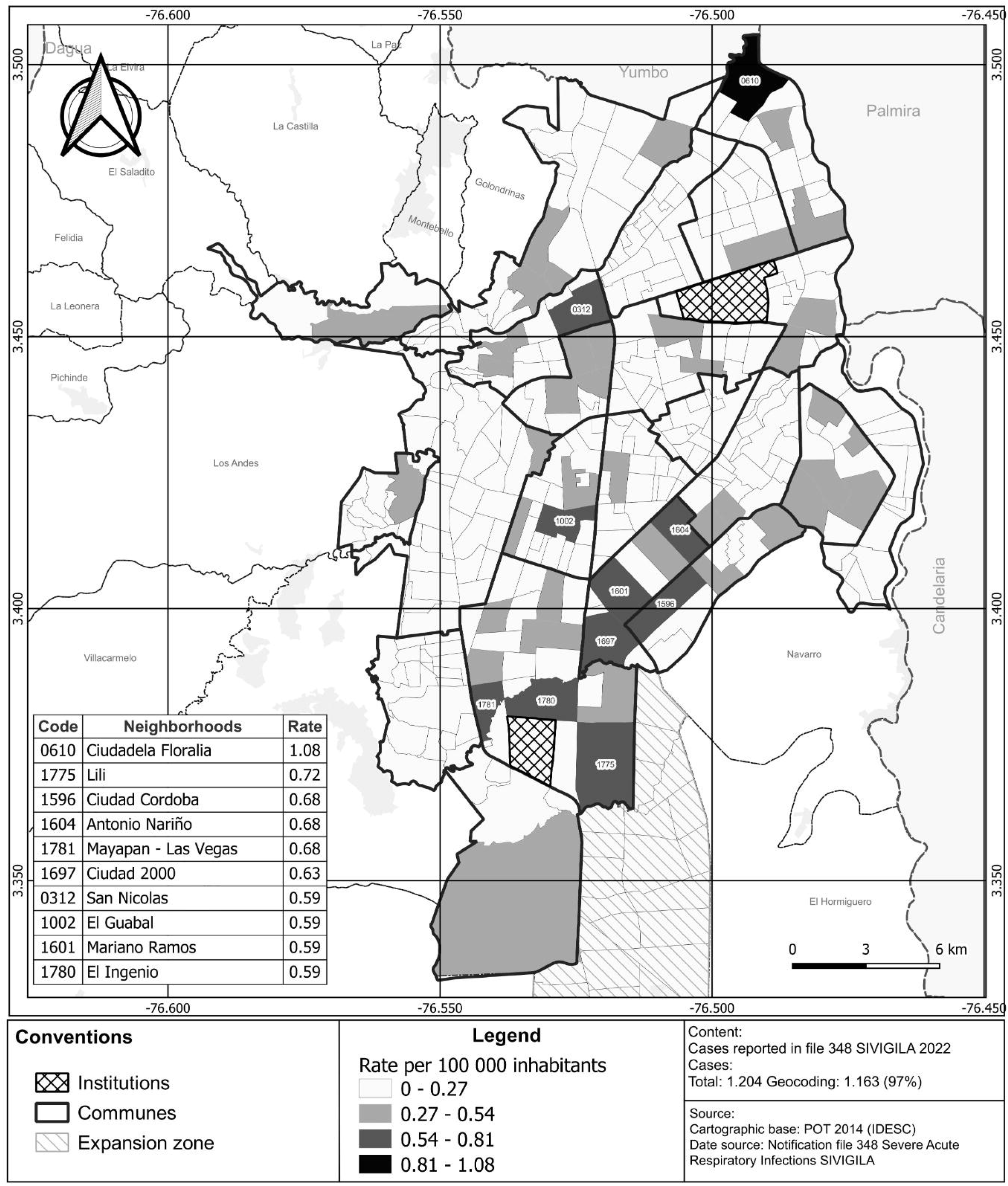

## Notes

### Competing Interest Statement

The authors have declared no competing interest.

### Funding Statement

This study did not receive any funding

### Author Declarations

Filing No.: 202441450100041674 Date: 2024-12-11 TRD: 4145.010.14.12.187.004167 Parent Filing No.: 202441450100041674 The Research Ethics Committee of the Health Secretariat of the Special District of Santiago de Cali reviewed the research project titled: Sociodemographic Determinants of Respiratory Failure in Patients Hospitalized with SARI in Cali, Colombia. After evaluating the documents submitted by the researcher, the committee considers the project approved for implementation. The project's risk level is classified as no risk. Any modification to the project must be reviewed and approved by the committee. The researcher must also report any issues related to data collection or unforeseen events that could pose a risk to the subjects. Additionally, a progress report must be submitted every three months. The committee may revoke the approval of a research project in progress if it is determined that false data has been provided, additional risks not mentioned in the initial submission are present, or deficiencies in the conduct of the study are identified. The revocation of approval will take immediate effect and will be notified via email.

